# Lesion selection and external evaluation of clinical–imaging survival models after transarterial chemoembolization for hepatocellular carcinoma

**DOI:** 10.64898/2026.10.28.26366759

**Authors:** Zhongling Xu, Alaa Melek, Edward Castillo, Hairong Wang

## Abstract

**Background:** Pretreatment computed tomography (CT) describes tumor characteristics in patients receiving transarterial chemoembolization (TACE) for hepatocellular carcinoma (HCC), but whether measuring more tumors improves mortality predictions in a new cohort remains unclear. We assessed the added value of different lesion-selection scopes beyond shared clinical predictors.

**Methods:** We compared a model using five clinical variables with models adding CT measurements from the largest, two largest, or all retained annotated tumors. All models used the same regularized discrete-time survival estimator. Evaluation used repeated nested cross-validation in 228 WAW-TACE patients and frozen external application in 69 HCC-TACE-Seg patients, with 15 deaths by day 365. We assessed first-year probability error, discrimination, and calibration, followed by local recalibration using small external samples.

**Results:** Internally, adding CT-derived lesion features did not establish an improvement in prediction accuracy. Externally, the all-lesion model had the highest discrimination point estimates, but its advantage over the clinical-only model was uncertain. Its mortality probabilities were less accurate than those of the clinical-only model, with calibration estimates consistent with overly extreme predicted risks. Local recalibration reduced its median held-out prediction error at day 365, but the median adjusted error remained higher than that of the adjusted clinical model. Simple tumor-burden predictors also did not improve external probability accuracy over the clinical baseline.

**Conclusions:** In these cohorts, measurements from more annotated tumors did not yield more reliable first-year mortality probabilities. Comparisons of clinical–imaging models should evaluate external probability accuracy and calibration alongside patient ranking.

## 1 Background

Transarterial chemoembolization (TACE) is an established liver-directed therapy for selected patients with hepatocellular carcinoma (HCC), particularly within the heterogeneous intermediate-stage population [1]. Outcomes vary with liver function, tumor burden, and other clinical characteristics. Established prognostic systems use a few interpretable measurements. The albumin–bilirubin (ALBI) grade summarizes hepatic reserve; the hepatoma arterial-embolization prognostic (HAP) score combines albumin, bilirubin, alpha-fetoprotein (AFP), and dominant tumor size; modified HAP-II (mHAP-II) adds tumor count; and Six-and-Twelve combines tumor diameter with tumor number [2–5]. Pretreatment computed tomography (CT) also describes tumor shape, attenuation, and differences among tumors. Whether these measurements improve survival-risk estimates beyond clinical variables, particularly in a new cohort, remains an important evaluation question.

Many imaging-based TACE studies predict early objective response using tumor features, surrounding tissue, clinical variables, or multiple contrast phases [6–13]. Modified Response Evaluation Criteria in Solid Tumors (mRECIST) assesses viable enhancing tumor after locoregional therapy [14]. Other studies address TACE refractoriness or suitability [15, 16]. These endpoints differ from mortality: overall survival also reflects liver function, disease burden, subsequent treatment, and other clinical events. Response-prediction results therefore do not establish whether pretreatment CT improves survival-risk estimates.

Survival models after TACE include neural estimators using clinical and conventional tumor-burden variables [17], clinical–radiomic models [18, 19], deep-learning image features [20], and multi-organ radiomics [21]. Multiphase measurements provide another source of imaging information [10, 11, 18, 22]. Reviews report variation in populations, regions of interest, endpoints, and validation methods [23–26]. Cross-cohort evaluation has also been used for response prediction, including training on HCC-TACE-Seg and testing on WAW-TACE [27].

Lesion selection determines which imaging information enters a patient-level model. Clinical studies show that the number of target lesions can affect response assessment and its association with survival [28, 29]. Survival-radiomics studies have used the largest lesion [18] or delineated all lesions for texture analysis [19]. Using additional tumors could capture their total volume and variation in morphology or attenuation, while increasing sensitivity to small segmented regions and annotation granularity. Comparing the largest, two largest, and all annotated tumors under the same clinical inputs and estimator directly tests these competing considerations.

A contemporaneous study using WAW-TACE and HCC-TACE-Seg reported limited incremental value of CT radiomics and limited cross-cohort discrimination of frozen imaging models [30]. We examine first-year mortality using shared clinical predictors, a common lesion-aggregation scheme, and frozen clinical-only and combined clinical–imaging models. A model can rank patients well yet give inaccurate death probabilities in a new cohort. Brier scores and calibration therefore complement discrimination when assessing external predictions [31–33].

We developed the models in WAW-TACE [34] and evaluated them in HCC-TACE-Seg [35]. This study makes three contributions:

- **A controlled comparison of lesion representations**. A shared aggregation scheme compares the largest, two largest, and all retained annotated tumors against a clinical-only baseline, with the clinical predictors, survival estimator, and patient-level splits held constant.
- **Frozen external evaluation of both ranking and probability accuracy**. Clinical-only and combined models are applied without retraining or recalibration, allowing discrimination, prediction error, and calibration to be assessed separately.
- **Evaluation of limited-sample local adaptation**. Small external calibration samples quantify the feasibility and held-out accuracy of risk adjustment. Tumor-burden and fixed-capacity prioritization analyses provide complementary comparisons of input complexity and patient ranking.

## 2 Methods

### 2.1 Study Design, Data Sources, and Outcome Definition

This retrospective prognostic modeling study used two public cohorts of patients with HCC treated with TACE. WAW-TACE was assigned to model development and internal validation, and HCC-TACE-Seg was reserved for external evaluation [34, 35]. Only information available before the first TACE was used. The target was patient-level mortality risk during the following 365 days; the target population comprised patients already selected for TACE. Figure 1 summarizes the data processing, model comparisons, and validation procedures. Additional file 1 provides supplementary tables, figures, and methods.

**Figure 1.**
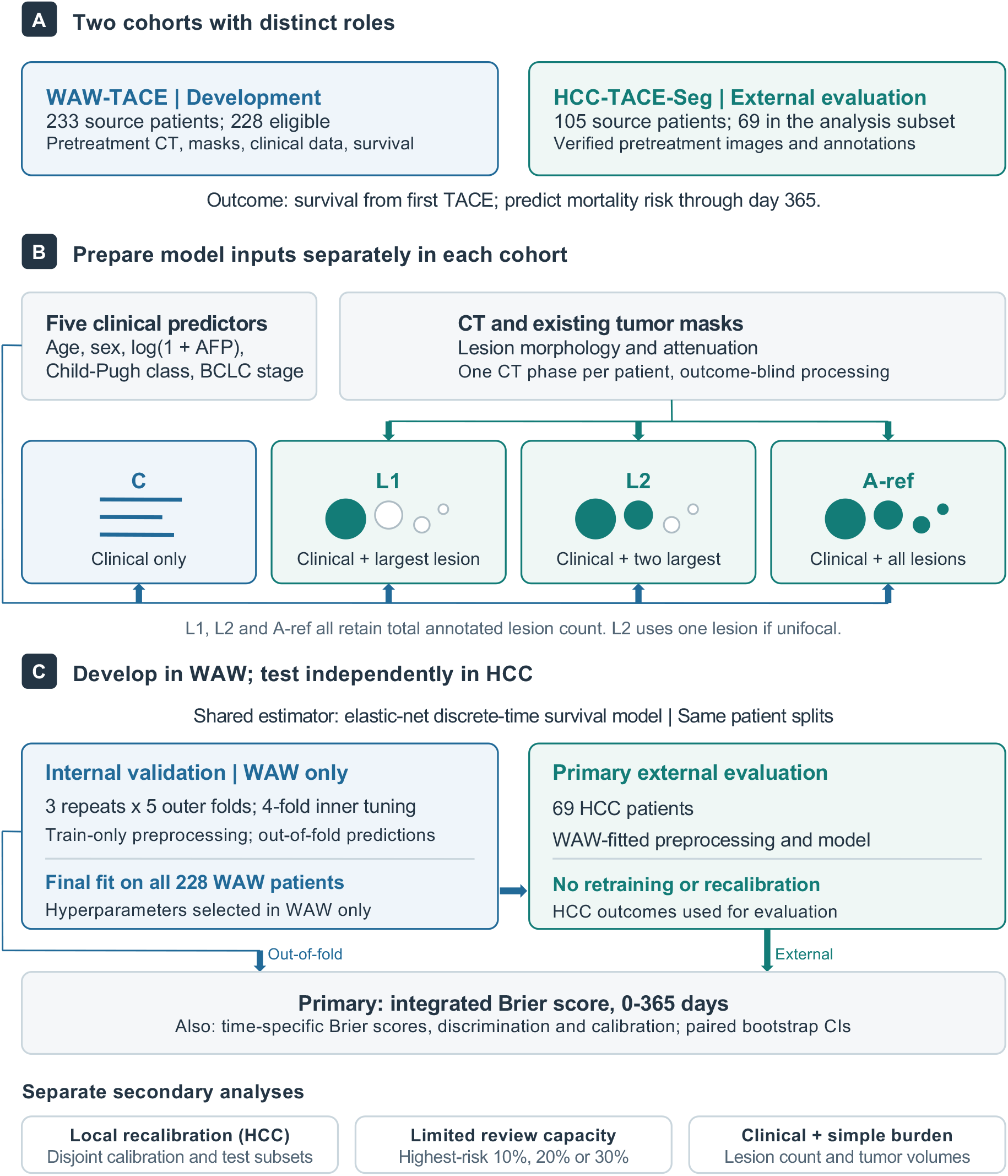
Study workflow. The clinical-only model (C) used five clinical predictors; L1, L2, and A-ref added CT features from the largest lesion, the two largest lesions, or all annotated lesions, respectively. All models used the same estimator and patient splits, and image processing was outcome-blind. Models were developed in WAW-TACE. Primary external evaluation in HCC-TACE-Seg used these models without retraining or recalibration.

WAW-TACE contains 233 patients, multiphase pretreatment CT, clinical variables, survival outcomes, and 377 manually annotated tumor masks. Patients were treated at the Central Clinical Hospital of the Medical University of Warsaw, Poland, between May 2016 and April 2021 [34]. Its CT scans cover precontrast/native (NC), late arterial (A), portal venous (PV), and delayed phases. Each patient’s tumor masks are associated with one of these phases.

The HCC-TACE-Seg source collection contains 105 patients with pre- and post-TACE imaging, clinical variables, curated tumor and liver masks, and time-to-event outcomes. Patients were treated at The University of Texas MD Anderson Cancer Center, USA, between November 2002 and June 2012 [35]. The external analysis used an existing subset of 69 patients with verified pretreatment NC, A, and PV images and available tumor annotations. Availability of all three phases was a cohort-assembly criterion; imaging predictors for this study were extracted only from the mask-associated PV image (Section 2.2).

Eligibility was assessed in the 233-patient WAW-TACE cohort and the assembled 69-patient HCC-TACE-Seg subset. Patients required at least one usable pretreatment tumor annotation, a positive overall-survival time, and a valid death or censoring indicator. Image and mask checks were performed without access to survival outcomes. Five WAW-TACE patients were excluded during image QC: one because CT–mask geometry could not be reconciled and four because less than 80% of lesion voxels fell within the liver-support mask. The analytic cohorts comprised 228 WAW-TACE patients and all 69 assessed HCC-TACE-Seg patients (Table 1; Additional file 1: Table S4).

**Table 1.** Cohort eligibility and selected analytic-cohort characteristics.

| **Panel A: Cohort eligibility and annotation counts** |  |  |
| --- | --- | --- |
| Item | WAW-TACE | HCC-TACE-Seg |
| Cohort role | Development | External evaluation |
| Patients in the source cohort | 233 | 105 |
| Patients assessed for this analysis | 233 | 69 |
| Patients not retained after eligibility assessment | 5 | 0 |
| Model-eligible patients | 228 | 69 |
| Lesion instances/components before image QC | 377 | 122 |
| Retained tumor instances/components | 372 | 122 |

| **Panel B: Selected analytic-cohort characteristics** |  |  |
| --- | --- | --- |
| Characteristic | WAW-TACE<br>(*n* = 228) | HCC-TACE-Seg<br>(*n* = 69) |
| Age, years | 66 [60, 73] | 71 [60, 76] |
| Patients with multiple annotated lesions, *n* (%) | 85 (37.3) | 14 (20.3) |
| Total tumor volume, mL | 50.1 [18.9, 113.0] | 141.2 [39.0, 323.5] |
| Deaths by day 365, *n* (%) | 60 (26.3) | 15 (21.7) |

*Note.* Eligibility assessment in HCC-TACE-Seg was performed within the assembled 69-patient subset of the 105-patient source cohort. Panel A lesion counts refer to the assessed patients: individual lesion masks in WAW-TACE and connected components of patient-level tumor masks in HCC-TACE-Seg. Panel B describes the model-eligible patients; values are median [Q1, Q3] or *n* (%), as indicated. Percentages use the analytic-cohort denominators. Patients with multiple annotated lesions had at least two retained annotations.

The primary endpoint was overall survival from the first TACE. WAW-TACE recorded survival time in days. HCC-TACE-Seg recorded overall survival in weeks, which was multiplied by seven. For survival analysis, a recorded death counted as an event. Otherwise, the patient was right-censored at the last documented follow-up. Because survival after that time was unknown, patients censored before a prediction time point were not counted as known survivors at that point. Post-TACE imaging, radiological response, progression variables, subsequent treatment, and future TACE count were excluded from all predictors.

The shared clinical predictors were age, sex, alpha-fetoprotein (AFP), Child–Pugh class, and ordinal Barcelona Clinic Liver Cancer (BCLC) stage. Age was entered in years, and sex was encoded as 0 for male and 1 for female. AFP, recorded in ng/mL, was transformed as log(1 + AFP) to reduce the influence of very high values while allowing a recorded value of zero.

Child–Pugh classes A, B, and C were encoded as 1, 2, and 3, respectively; these values represent ordered classes, not the original Child–Pugh score. BCLC stages 0/A, B, C, and D were encoded as 0, 1, 2, and 3, respectively, with stage 0 grouped with stage A to match the shared cohort coding. The two ordered predictors were entered numerically rather than as separate category indicators. Cohort-specific source fields and their conversion to these shared encodings are listed in Additional file 1: Table S10. Missing values and differences in scale were handled within the training data, as described in Section 2.3. Cohort-specific variables, including albumin and bilirubin in WAW-TACE and performance status in HCC-TACE-Seg, were excluded from the prediction models.

Published scores including ALBI, HAP, mHAP-II, and Six-and-Twelve were not used as fixed benchmarks because their required components and units could not be reconstructed consistently across cohorts [2–5] (Additional file 1: Table S3).

Table 1 summarizes cohort eligibility and selected characteristics of the analytic samples, following reporting guidance for clinical prediction models and medical-imaging AI [36, 37]. Data provenance and analysis records are described in Additional file 1: Section S2 (*Data Provenance and Analysis Records*).

### 2.2 Image Processing and Feature Construction

Image processing converted each patient’s CT and tumor annotations into a fixed set of patient-level predictors. The primary analysis used only the mask-associated reference CT, not multiphase features. For WAW-TACE, the reference image was the official mask-associated CT phase. For HCC-TACE-Seg, it was the verified pretreatment PV image on which the retained tumor and liver masks were defined. Masks were aligned to the reference image grid only when their spatial metadata confirmed the same physical coordinate frame.

WAW-TACE provides one binary tumor-mask file for each annotated lesion, so the masks were preserved as distinct lesion instances. By contrast, HCC-TACE-Seg provides a single patient-level tumor mask, which was separated into lesion instances using three-dimensional face-connected components (6-connectivity). Every nonempty component was retained; no minimum-volume threshold or distance-based merging was applied. Lesions were ranked by decreasing physical volume, with deterministic tie-breaking.

Before feature extraction, each lesion mask had to be spatially compatible with its reference CT. In WAW-TACE, at least 80% of lesion voxels also had to fall within the provided liver segment. In HCC-TACE-Seg, the same threshold used the union of liver and tumor masks, which included the tumor mask itself and therefore did not independently assess liver overlap. CT values were clipped to [*−* 200, 400] Hounsfield units (HU). Lesion definition, geometry reconciliation, quality control, and feature extraction were completed before survival outcomes were joined.

For each retained lesion, we calculated seven features describing size, shape, and attenuation (Table 2). The bounding-box diagonal was a three-dimensional size descriptor, not a RECIST diameter. Attenuation was summarized relative to a reference liver region on the same CT, defined as voxels in the provided liver mask after excluding the lesion being measured. The lesion-to-reference-liver difference was

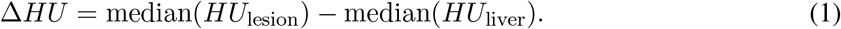

**Table 2.** Imaging features used to construct patient-level model inputs.

| Feature | Unit | Definition |
| --- | --- | --- |
| *Lesion-level features, each reduced to five patient-level summaries* |  |  |
| Volume | mL | Mask voxel count multiplied by physical voxel volume. |
| Bounding-box diagonal | mm | Diagonal of the three-dimensional physical bounding box. |
| Surface-to-volume ratio | $\text{mm}^{-1}$ | Voxel-face surface area divided by lesion volume in $\text{mm}^3$. |
| Sphericity | — | Surface area of a sphere with the same volume divided by lesion surface area. |
| Relative attenuation | HU | Median lesion HU minus median reference-liver HU. |
| Attenuation interquartile range | HU | Difference between the 75th and 25th percentiles of lesion HU. |
| High-attenuation fraction | — | Fraction of lesion voxels at least 20 HU above the median reference-liver HU. |
| *Patient-level burden predictors, entered directly* |  |  |
| Total annotated lesion count | Count | Number of retained lesions, identical across L1, L2, and A-ref. |
| Included lesion count | Count | Number of lesions selected by the representation. |
| Selected tumor volume | mL | Sum of the selected lesion volumes. |
| Small-lesion volume fraction | — | Selected volume from lesions smaller than 1 mL divided by total selected volume. |

*Note.* For each lesion-level feature, the five summaries are the largest-lesion value, volume-weighted mean, maximum, unweighted standard deviation, and range. For a single selected lesion, the first three equal its feature value and the last two are zero. “—” denotes a dimensionless quantity; fractions are on a 0–1 scale. HU denotes Hounsfield units.

Geometry checks, liver-support definitions, tie-breaking, feature formulas, and the reference-liver definition are given in Additional file 1: Section S3.2 (*Lesion geometry and attenuation*).

The largest-one representation (L1) selected the largest lesion; the largest-two representation (L2) selected the two largest lesions, or the single available lesion for a unifocal patient; and the all-lesion reference-CT representation (A-ref) selected every retained annotated lesion. For each of the seven features, we retained its value in the largest selected lesion. We also computed its volume-weighted mean, maximum, unweighted standard deviation, and range across the selected lesions, giving five patient-level summaries per feature. These summaries describe both representative lesion characteristics and differences between lesions.

Four burden predictors were added to each representation (Table 2). The total annotated lesion count was retained in all three imaging representations, even when only one or two lesions supplied the other imaging features. Figure 2A summarizes how lesion selection and aggregation produce patient-level imaging predictors, which were combined with the five clinical predictors defined in Section 2.1.

**Figure 2.**
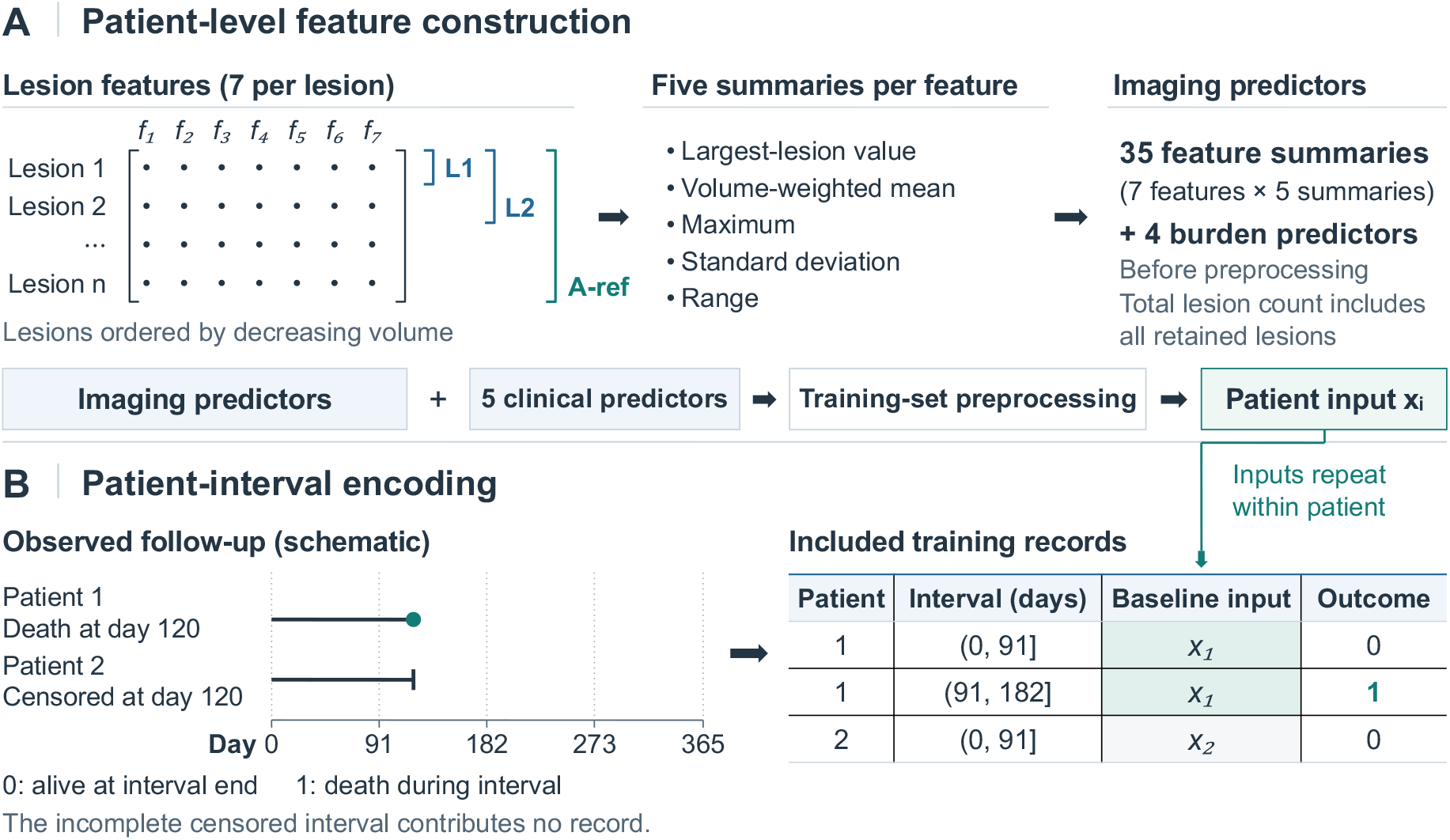
Model input construction and patient–interval encoding. (A) Lesion-selection scopes and the shared aggregation scheme. The 35 summaries and four burden predictors are counted before preprocessing; L2 uses the available lesion for a unifocal patient. (B) Schematic follow-up and the resulting training records, with the patient’s baseline input repeated across intervals. Outcomes 0 and 1 denote survival through an interval and death within it, respectively. The incomplete censored interval contributes no record.

### 2.3 Model Development and Evaluation Protocol

To isolate the contribution of lesion-selection scope while limiting overfitting, we compared four models using the same regularized estimator and patient splits. The clinical-only model (C) used the five shared clinical predictors defined in Section 2.1. L1, L2, and A-ref added the imaging predictors constructed in Section 2.2; A-ref was the primary imaging model. The three imaging models differed only in the lesions used to calculate their imaging summaries. We used prespecified clinical and lesion-summary features to limit model complexity in the 228-patient development cohort.

#### Model specification

We used a discrete-time survival model to predict the probability of death in successive time intervals. The first 365 days after TACE were divided into four intervals ending at days 91, 182, 273, and 365. Each patient’s follow-up was expanded into one binary record per usable interval. A record was labeled 0 when the patient was known to be alive at the end of the interval and 1 when death occurred during that interval. An incomplete censored interval contributed no record (Figure 2B). Each record contained the preprocessed baseline vector **x**_*i*_, the interval indicator, and the binary death label. The vector combined the clinical predictors with the imaging predictors in Table 2 for L1, L2, and A-ref, and was unchanged across a patient’s records. We fitted a single *pooled logistic regression* model to all usable patient–interval records in the training set. The conditional probability of death in each interval was modeled as

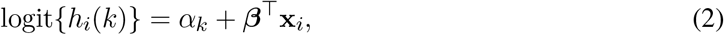

and converted to cumulative mortality risk by

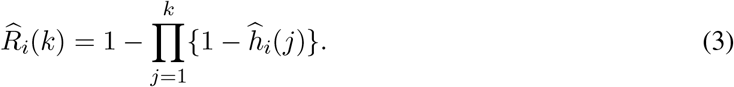

Here, *h*_*i*_(*k*) is the probability that patient *i* dies during interval *k*, conditional on being alive at its start, and logit(*p*) = log{ *p/*(1− *p*) }. The fitted predictor coefficients ***β*** are shared across intervals, whereas the intercepts *α*_*k*_ allow baseline death risk to differ across the four periods. 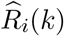 is predicted cumulative mortality risk through interval *k*.

Elastic-net regularization combines an L1 penalty, which can set coefficients to zero, with an L2 penalty, which shrinks coefficients and helps stabilize correlated predictors. Writing *α*_*k*_ = *a* + *d*_*k*_, with *d*_1_ = 0, separates the common intercept *a* from three interval-indicator coefficients *d*_2_, *d*_3_, *d*_4_. Let ***θ*** contain these three coefficients and ***β***. The fitted parameters minimized

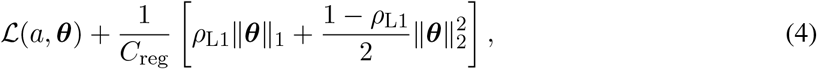

where *L* is the sum of binary negative log-likelihoods over the pooled records, ∥***θ***∥ _1_ is the sum of absolute coefficients, and 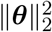 is the sum of squared coefficients. Both the predictor coefficients and interval-indicator coefficients were penalized; the common intercept *a* was not. The full design matrix and record-level likelihood are given in Additional file 1: Section S3.3 (*Pooled-logistic design and penalized likelihood*).

#### Training and validation

Internal performance was estimated in WAW-TACE using three different random patient-level partitions into five folds. Each partition defined a complete five-fold outer cross-validation run: four folds were used for model development and the remaining fold was held out for evaluation, rotating until every patient had been held out once. Within each outer training set, four-fold inner cross-validation selected the elastic-net parameters with the lowest mean validation integrated Brier score (IBS), the primary measure of probability error over 0–365 days described below.

Within each cross-validation training set, predictors missing for all patients were removed. Other missing values were replaced by the corresponding training-set median, after which predictors that were constant in the training data were removed. Each retained predictor was then centered and scaled using the mean and standard deviation estimated from that training set. The same fitted transformations were applied to the corresponding validation or held-out patients. The tuning grid used inverse regularization strengths *C*_reg_ ∈ { 0.01, 0.1, 1, 10}, for which smaller values impose stronger regularization, and L1 mixing proportions *ρ*_L1_ ∈ { 0, 0.25, 0.5, 0.75, 1 }, where 0 denotes pure L2 regularization and 1 denotes pure L1 regularization.

All preprocessing and parameter selection used only the training patients in each split. The three outer partitions produced three sets of four time-point risk predictions for each patient, each from a model trained without that patient. These predictions were averaged for the primary internal evaluation.

For external evaluation, four-fold cross-validation on all 228 eligible WAW-TACE patients selected hyperparameters using the same grid and IBS criterion. Each model was then fitted to the full development cohort. The resulting preprocessing transformations and model coefficients were applied to HCC-TACE-Seg without refitting or recalibration. Local recalibration was examined separately in Section 2.4. Selected hyperparameters and clinical-predictor missingness are summarized in Additional file 1: Table S2.

#### Performance evaluation

The primary measure was the inverse-probability-of-censoring-weighted (IPCW) IBS, which summarizes mortality-probability accuracy over 0–365 days while accounting for incomplete follow-up.

At a given time *t*, the Brier score is the mean squared difference between predicted mortality risk and observed death status. For *n* evaluated patients, the IPCW Brier score and its time-integrated summary are

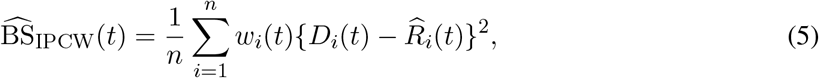

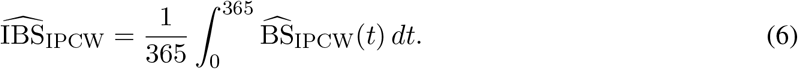

Here, 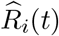 is predicted mortality risk by day *t*, and *D*_*i*_(*t*) = 1 for a documented death by *t* and 0 otherwise. The weight *w*_*i*_(*t*) is zero for patients censored by *t*, whose status at that time is unknown. For a documented death, the weight is the inverse estimated probability of remaining uncensored until the death time; for a patient known to survive beyond *t*, it is the inverse probability of remaining uncensored until *t*. These probabilities were estimated by Kaplan–Meier analysis of censoring in the evaluated sample [38]. IPCW therefore gives more weight to observable outcomes that represent patients with incomplete follow-up. The Brier score describes accuracy at one time point, whereas IBS summarizes accuracy across the follow-up period; lower values are better for both [31, 32].

For computation, we used the four interval-end risk predictions and approximated IBS by trapezoidal integration on days 0, 91, 182, 273, and 365, setting the day-0 Brier score to zero. Further calculation details are given in Additional file 1: Section S3.4 (*Numerical integration, calibration, and uncertainty*).

Secondary measures were the IPCW Brier score, IPCW-weighted time-dependent area under the receiver operating characteristic curve (AUROC), and average precision at 91, 182, 273, and 365 days, together with Harrell’s concordance index based on 365-day risk [39]. Time-dependent AUROC measures how well predicted risk ranks patients who die by a given time above those who are known to be alive beyond that time, while average precision summarizes the corresponding precision–recall performance. Harrell’s concordance index measures how often a patient who dies earlier receives a higher predicted risk; higher values indicate better ranking for all three measures.

Calibration at 365 days assessed agreement between predicted and observed mortality risk. We fitted an IPCW logistic regression of death status on the logit of predicted risk, estimating its intercept and slope jointly. Ideal calibration corresponds to an intercept of 0 and a slope of 1. A slope below 1 indicates that predicted risks are too extreme; a slope above 1 indicates that they are too similar to one another [33]. These calibration estimates were diagnostic and did not modify the primary predictions.

Ninety-five percent confidence intervals for the four primary models were calculated from 2,000 patient-level bootstrap replicates. For a cohort of *n* patients, each replicate was formed by drawing *n* patients at random with replacement. For model differences, the same resampled patient indices were used for both models and the difference was recalculated in each replicate. The prespecified internal comparisons were A-ref versus C, A-ref versus L1, and A-ref versus L2. The reporting analysis additionally estimated the remaining internal pairwise contrasts and all external pairwise contrasts. These intervals were not adjusted for multiple comparisons.

### 2.4 Secondary Analyses

#### Local recalibration

We examined whether small local calibration samples from HCC-TACE-Seg could improve predictions of cumulative mortality by day 365 from the clinical baseline and A-ref, the primary imaging model. We kept the original survival models fixed and fitted a separate recalibration mapping to their 365-day predictions. For each calibration sample size of 10, 20, or 40 patients, we used 200 fixed-seed random patient-level splits of the 69 HCC patients, holding out the remaining patients for evaluation. Within each calibration subset, we fitted an IPCW logistic regression using the logit of the original predicted risk as the predictor. The binary outcome was coded as 1 for death by day 365 and 0 for known survival beyond that day; patients with unknown 365-day status had zero IPCW weight. The recalibration model adjusted either its intercept alone, with the slope fixed at 1, or both its intercept and slope. Intercept-only adjustment required at least five deaths by day 365 and five patients known to survive beyond that day in the calibration subset. Adjusting both parameters required at least 10 of each.

The IPCW weights defined in Section 2.3 were estimated separately from the calibration and held-out patients. Calibration-subset weights were used to fit the adjustment; held-out-subset weights were used only to calculate evaluation scores. Feasibility was reported as the number of splits in which recalibration and evaluation could be completed, out of all 200 attempted (Additional file 1: Table S5). Only these splits contributed to performance summaries. We compared original and adjusted IPCW Brier scores at 365 days on the same held-out patients, reporting the median and interquartile range (IQR) of the paired differences across splits. These ranges describe split-to-split variation, not confidence intervals. Recalibration targeted only 365-day risk, so adjusted IBS over 0–365 days was not calculated.

#### Risk-based prioritization

To compare prioritization under a fixed review capacity, we ranked HCC patients using the original, uncalibrated 365-day predictions from C and A-ref. The review budget was the number of patients selected for further assessment. At the 10%, 20%, and 30% levels, each model selected its 7, 14, and 21 highest-risk patients, respectively, from the 69-patient cohort. We estimated the fraction of all deaths by day 365 captured in the selected group, with IPCW adjustment and 2,000 paired patient-bootstrap resamples. The 20% review budget was the principal comparison. This retrospective exercise represents prioritization for further clinical assessment, not the effect of that assessment on survival.

#### Tumor burden and cohort differences

In a subsequent secondary analysis, we evaluated a C+burden model that combined the five clinical predictors with annotated lesion count, total tumor volume, and largest-lesion volume. This model used the same preprocessing and tuning grid as the four primary models. It was evaluated in WAW by the original repeated nested procedure and then applied without refitting to HCC. Differences from C and A-ref used 1,000 paired patient-bootstrap resamples.

Separately, we described differences in clinical and image-feature distributions between the development and external cohorts using medians and standardized mean differences (Additional file 1: Table S8). These summaries used no survival outcomes and did not guide feature selection.

## 3 Results

### 3.1 Cohort Characteristics and Outcome Support

The development cohort comprised 228 WAW-TACE patients with 372 retained tumor instances; the external cohort comprised 69 HCC-TACE-Seg patients with 122 retained lesion components. At least two retained annotated lesions were present in 85 WAW patients (37.3%) and 14 HCC patients (20.3%) (Table 1; Additional file 1: Table S4). By day 365, 60 WAW patients had died and 168 were known to have survived beyond that day. In HCC, 15 had died, 53 were known survivors, and one had been censored before day 365.

The cohorts differed in disease stage, tumor burden, and reference CT phase. WAW included only BCLC stages 0/A and B, whereas 46 of the 69 HCC patients (66.7%) were at stages C or D. Median total tumor volume was 50.1 and 141.2 mL, respectively (Additional file 1: Table S8). WAW reference images included arterial, portal-venous, delayed, and native phases, whereas all HCC reference images were portal venous.

### 3.2 Internal Predictive Performance

The four primary models had closely spaced IBS estimates in repeated nested out-of-fold validation, ranging from 0.0934 to 0.0955 (Table 3A). A-ref had an IBS of 0.0951, compared with 0.0942 for the clinical baseline, C. The paired A-ref-minus-C difference was 0.0009 (95% CI, *−* 0.0047 to 0.0064). All six paired IBS intervals included zero, so the data did not establish an internal improvement in probability accuracy from adding the lesion measurements (Figure 3A; Additional file 1: Table S1).

**Table 3.** Internal validation and coefficient-frozen external performance.

A: WAW-TACE repeated nested out-of-fold predictions (n = 228)*
| Model | IBS (95% CI) | Brier<sub>365</sub> | AUROC<sub>365</sub> | Calibration slope |
| --- | --- | --- | --- | --- |
| C | 0.0942 [0.0753, 0.1156] | 0.1842 | 0.6529 | 0.744 |
| L1 | 0.0955 [0.0766, 0.1153] | 0.1797 | 0.6746 | 0.929 |
| L2 | 0.0934 [0.0728, 0.1152] | 0.1751 | 0.6704 | 1.073 |
| A-ref | 0.0951 [0.0762, 0.1150] | 0.1778 | 0.6721 | 1.041 |
| C+burden* | 0.0944 [0.0748, 0.1151] | 0.1738 | 0.6937 | 1.148 |

*Panel B: HCC-TACE-Seg frozen external predictions (n = 69)*
| Model | IBS (95% CI) | Brier<sub>365</sub> | AUROC<sub>365</sub> | Calibration slope |
| --- | --- | --- | --- | --- |
| C | 0.0631 [0.0435, 0.0867] | 0.1583 | 0.6351 | 0.886 |
| L1 | 0.1195 [0.0863, 0.1554] | 0.2342 | 0.6472 | 0.245 |
| L2 | 0.1651 [0.1231, 0.2118] | 0.2749 | 0.6889 | 0.315 |
| A-ref | 0.1276 [0.0940, 0.1618] | 0.2317 | 0.7265 | 0.467 |
| C+burden* | 0.0914 [0.0609, 0.1256] | 0.2060 | 0.6029 | 0.138 |

**Figure 3.**
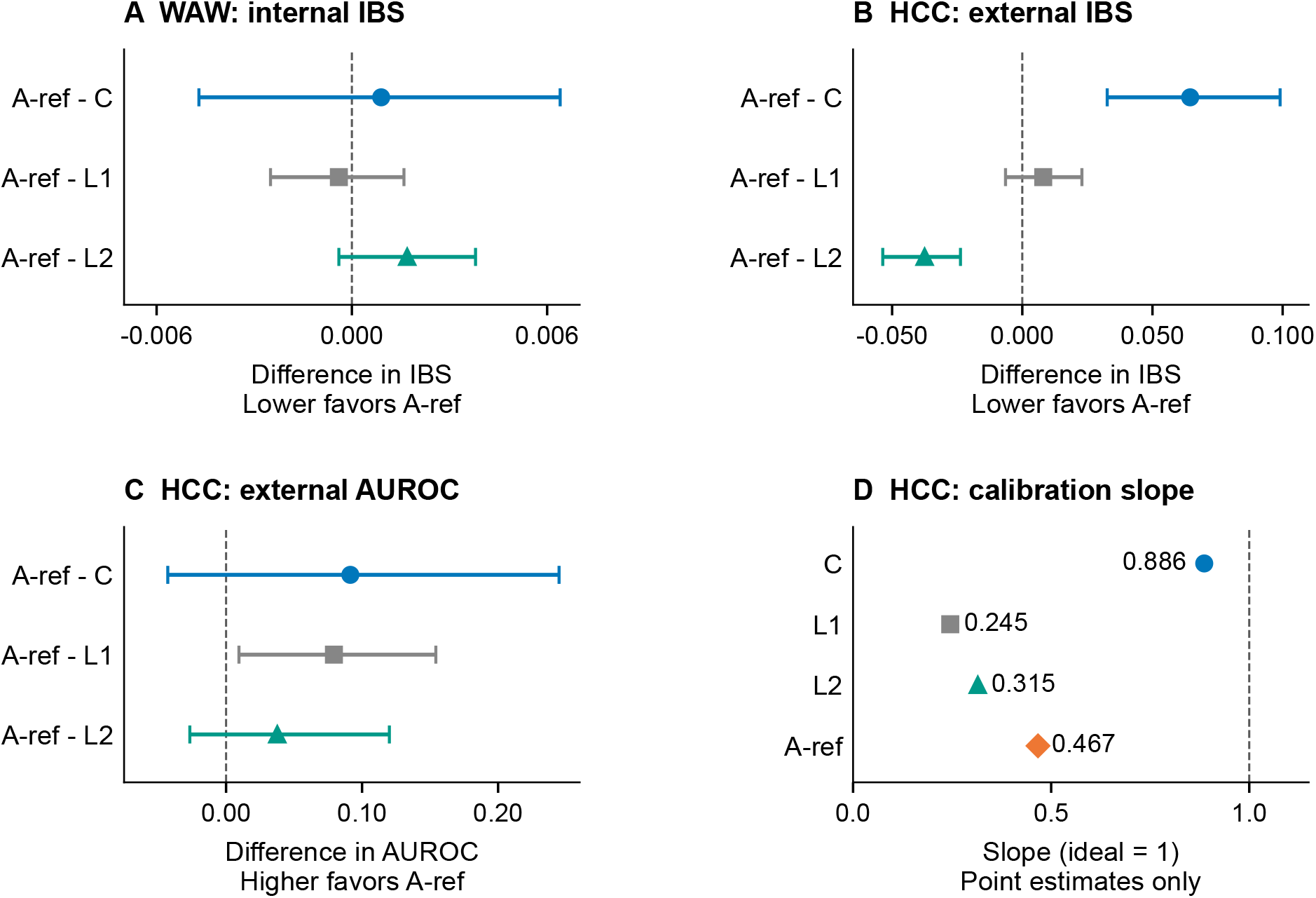
Paired performance contrasts and external calibration of the primary models. Panels A–C show A-ref minus each comparator: internal IBS (A), external IBS (B), and external AUROC at day 365 (C), with 95% paired patient-bootstrap intervals from 2,000 replicates. Negative differences favor A-ref for IBS; positive differences favor A-ref for AUROC. Panel D shows external calibration-slope point estimates, with 1 as the ideal reference. External results use the original, unadjusted predictions.

The imaging models had slightly higher 365-day AUROC point estimates than C (0.6704–0.6746 versus 0.6529), but none of the paired AUROC intervals excluded zero. Their calibration-slope point estimates were 0.929–1.073, compared with 0.744 for C. Additional performance measures, including day-365 average precision, C-index, and the calibration-model intercept, are reported in Additional file 1: Table S9.

### 3.3 External Risk Accuracy and Discrimination

In HCC-TACE-Seg, C had the lowest IBS (0.0631), compared with 0.1276 for A-ref, 0.1195 for L1, and 0.1651 for L2 (Table 3B). The paired A-ref-minus-C difference was 0.0645 (95% CI, 0.0326 to 0.0990), and paired IBS intervals favored C over all three imaging models (Additional file 1: Table S1). A-ref had lower IBS than L2, but its IBS comparison with L1 remained uncertain (Figure 3B).

A-ref had the highest point estimates for 365-day AUROC and C-index (0.7265 and 0.7072). Its AUROC difference from C was 0.0914, with a paired 95% CI of *−* 0.0429 to 0.2448. A-ref also had higher AUROC than L1, whereas the interval for A-ref versus L2 included zero (Figure 3C). C-index comparisons followed the same pattern (Additional file 1: Table S1). A-ref’s discrimination advantage over L1 was not accompanied by a clear reduction in probability error.

The external calibration-slope point estimates were below 1 for all four models: 0.886 for C, 0.467 for A-ref, 0.245 for L1, and 0.315 for L2 (Figure 3D). The lower slopes for the imaging models were consistent with predicted risks that were too extreme relative to observed outcomes. These diagnostic estimates did not alter the frozen predictions; local adjustment was evaluated separately below.

#### Secondary tumor-burden comparison

C+burden had a lower WAW 365-day Brier score than C (0.1738 versus 0.1842), but their IBS estimates were similar (0.0944 versus 0.0942). Externally, C+burden’s IBS of 0.0914 was higher than C’s (0.0631) and lower than A-ref’s (0.1276). Its AUROC was also lower than A-ref’s (0.6029 versus 0.7265). The simple burden model therefore did not improve external risk accuracy over C, despite lower error than A-ref (Additional file 1: Table S7).

### 3.4 Local Recalibration

Local recalibration was limited by the number of observed deaths. Under the prespecified support requirements, 6, 84, and 200 of the 200 splits supported intercept-only adjustment with calibration samples of 10, 20, and 40 patients, respectively. The supported splits were identical for C and A-ref. Intercept-and-slope adjustment was supported in no splits at 10 or 20 patients and in 67 splits at 40 patients (Figure 4A; Additional file 1: Table S5).

**Figure 4.**
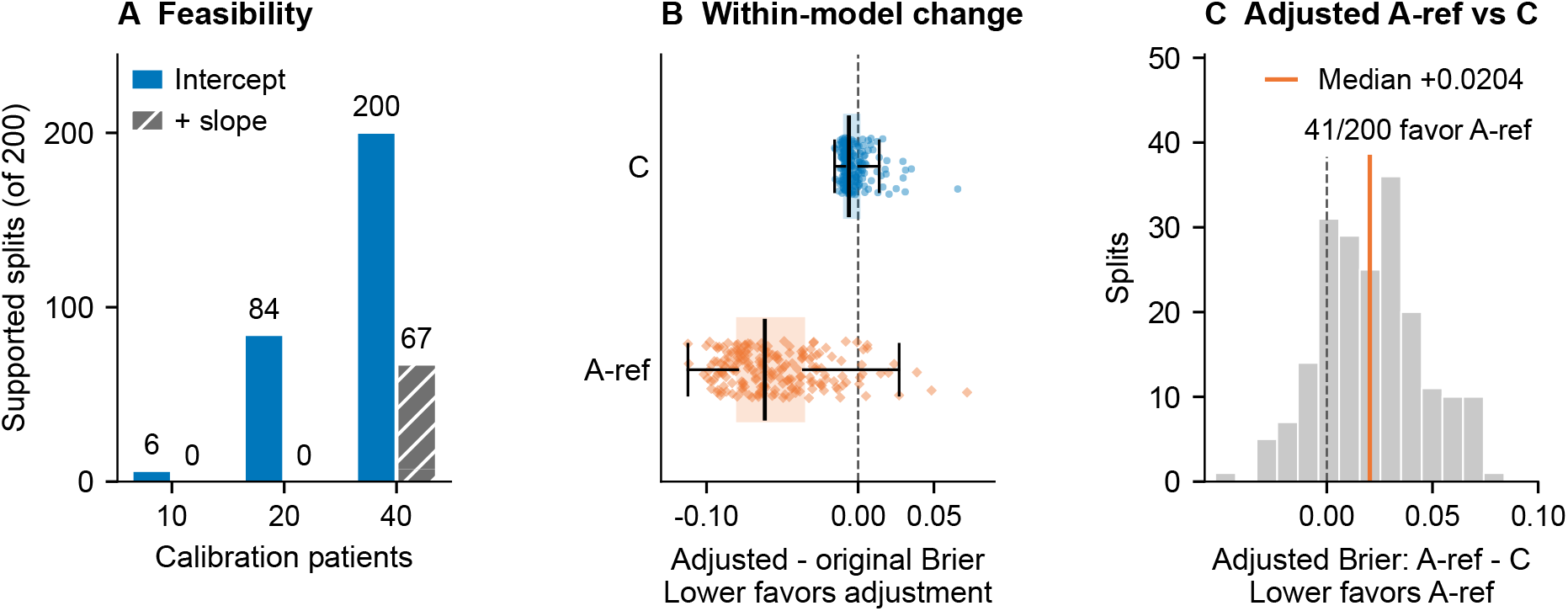
Feasibility and held-out error changes from local recalibration. Panel A counts supported splits at each calibration sample size; C and A-ref had identical support. Panels B and C use all 200 matched splits with 40 calibration and 29 evaluation patients. Panel B shows within-model changes after intercept adjustment; panel C shows adjusted A-ref minus adjusted C on the same held-out patients. Negative differences favor adjustment in B and A-ref in C. In B, points represent splits and boxes show medians and IQRs; whiskers extend to 1.5 IQR. The distributions summarize repeated splits within the same 69-patient cohort.

With 40 calibration patients and 29 held-out patients per split, intercept-only adjustment reduced A-ref’s 365-day Brier score relative to its original predictions (median paired change, *−*0.0616; split IQR, *−*0.0794 to *−*0.0361). The corresponding change for C was smaller (*−*0.0061; IQR, *−*0.0088 to 0.0004; Figure 4B). A-ref’s median adjusted Brier score remained higher than that of adjusted C (0.1727 versus 0.1491; Table 4). On matched splits, adjusted A-ref minus adjusted C had a median difference of 0.0204 (IQR, 0.0031 to 0.0366), and A-ref had lower error in 41 of 200 splits (Figure 4C).

**Table 4.**
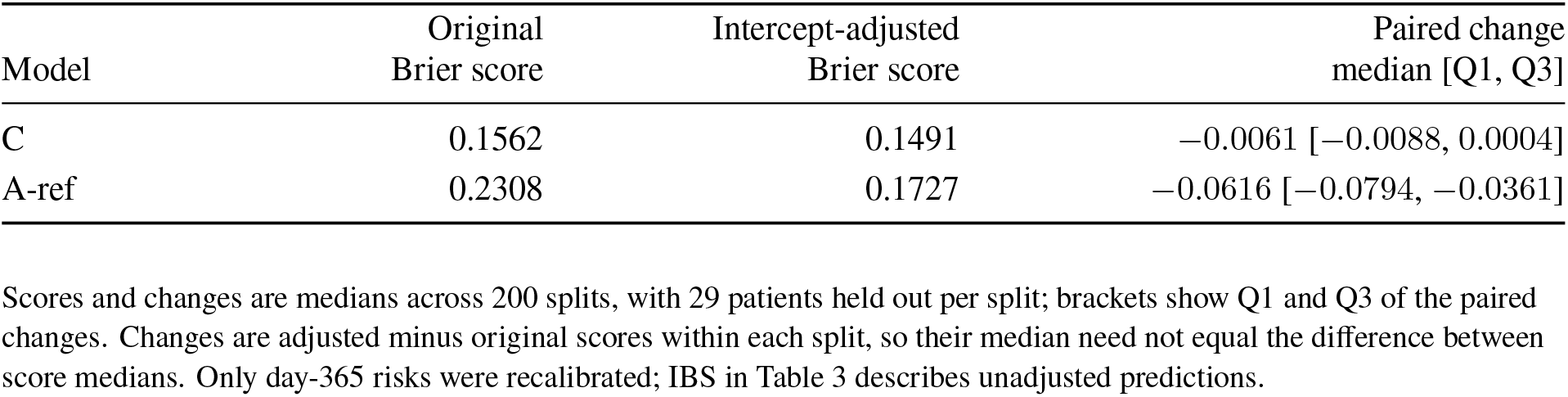
Held-out 365-day Brier scores after intercept-only recalibration with 40 HCC-TACE-Seg patients.

| Model | Original Brier score | Intercept-adjusted Brier score | Paired change median [Q1, Q3] |
| --- | --- | --- | --- |
| C | 0.1562 | 0.1491 | $-0.0061$ [$-0.0088$, $0.0004$] |
| A-ref | 0.2308 | 0.1727 | $-0.0616$ [$-0.0794$, $-0.0361$] |

Among the 67 splits that supported intercept-and-slope adjustment with 40 calibration patients, A-ref’s median Brier-score change relative to its original predictions was *−* 0.0880. Adjusted A-ref minus adjusted C had a median difference of 0.0071 (IQR, *−*0.0066 to *−*0.0155), with lower error for A-ref in 24 of the 67 matched splits (Additional file 1: Table S5). The two adjustment methods were summarized over different sets of supported splits, so these results do not provide a matched comparison between them.

### 3.5 Risk-Based Prioritization

When review was limited to the 14 HCC patients with the highest original predicted risk (20% of the cohort), C identified 7 of the 15 observed deaths by day 365, whereas A-ref identified 5. IPCW-adjusted event capture was 46.5% for C and 33.3% for A-ref. The paired A-ref-minus-C difference was *−* 13.2 percentage points (95% CI, *−* 25.0 to 18.8), so the data did not establish an A-ref advantage at this review capacity. At the 10% and 30% capacities, the models captured the same numbers of observed deaths (3 and 8, respectively), with similar IPCW-adjusted capture (Additional file 1: Table S6; Additional file 1: Figure S2).

## 4 Discussion

### 4.1 Principal findings and related evidence

None of the three lesion representations established an improvement in first-year survival-risk accuracy over the clinical baseline in WAW-TACE. In HCC-TACE-Seg, A-ref had the highest discrimination point estimates, but its discrimination advantage over C remained uncertain and its probability error was greater. Its external calibration slope was consistent with risks that were too extreme for the external cohort [33].

Lesion count and tumor burden contribute to established prognostic systems [3–5], which motivated testing whether morphology, attenuation, and variation among lesions added information to shared clinical variables. The C+burden comparison showed that simpler imaging-derived predictors also did not improve external accuracy over C (Additional file 1: Table S7). A-ref’s higher external AUROC than C+burden was accompanied by greater probability error, so the more detailed lesion representation did not offer a uniform performance advantage. Clinical–imaging fusion, multiphase radiomics, and deep learning have produced promising response or survival results in individual cohorts [8, 13, 18, 20]. Systematic reviews note that retrospective designs, heterogeneous regions of interest, inconsistent endpoints, and limited external validation constrain generalization [23–25]. Here, the frozen external comparison exposed an accuracy deficit that discrimination point estimates alone would not convey.

A contemporaneous analysis of the same public data sources also found no consistent incremental survival-prediction value from CT radiomics [30]. Its frozen external evaluation assessed an imaging score, while clinical combinations and baseline survival functions were fitted within the target cohort. Our external evaluation freezes the complete clinical-only and clinical–imaging models and assesses their first-year mortality probabilities. The controlled lesion-scope comparison and fixed-size local recalibration analyses provide complementary evidence on input construction and adaptation. Because the studies draw on the same source populations, their agreement reflects complementary analyses of overlapping data rather than replication in additional patients.

Differences in image acquisition, annotation, and case mix could contribute to the external error. WAW-TACE used the phase associated with each tumor mask, whereas the HCC-TACE-Seg reference was consistently PV. The cohorts also differed in stage and tumor volume (Additional file 1: Table S8). Because BCLC stage was modeled numerically, external predictions for stages C and D involved extrapolation beyond the development cohort’s observed stage range. Identical feature definitions therefore need not imply comparable feature distributions or relationships with outcomes. All-lesion aggregation may amplify variability because it includes small components, extreme values, and dispersion terms that depend on annotation granularity. These are possible contributors, rather than identified causes of the calibration error.

### 4.2 Local recalibration and risk-based prioritization

Intercept adjustment reduced A-ref’s median held-out Brier score relative to its original predictions, but its median matched-split error remained higher than that of adjusted C. An intercept adjustment changes the overall risk level without changing ranking or correcting an overly steep risk gradient.

At a fixed 20% review capacity, A-ref showed no established advantage over C in event capture despite its higher AUROC point estimate (Additional file 1: Table S6). AUROC evaluates ranking across the risk distribution, whereas fixed-capacity review evaluates only the highest-risk patients. A higher AUROC therefore need not yield greater event capture within the top 20%.

### 4.3 Strengths and limitations

Holding the estimator, clinical variables, and patient splits constant made the lesion representations comparable within each cohort. Outcome-blind lesion construction limited outcome-driven input selection, and nested internal validation and frozen external application kept model fitting separate from held-out evaluation.

Both cohorts were retrospective, and the external analysis included only 69 patients from an assembled triphasic subset, with 15 deaths by day 365. Selection of this subset may limit generalizability to the full HCC-TACE-Seg collection. Several paired discrimination intervals were wide. Only 14 patients (20.3%) had multiple retained annotated lesions; L1, L2, and A-ref selected the same lesion in the remaining patients. The external comparison therefore provides limited evidence about the value of broader lesion coverage in patients with multiple annotated lesions.

The conclusions apply to the compact reference-phase features tested here, with CT phase varying between cohorts, rather than harmonized multiphase or learned representations. “All lesions” denotes retained annotated components, which may differ from clinical lesion definitions or omit visible tumors. Available fields and measurement definitions did not permit exact reconstruction of published clinical scores.

Only 67 of the 40-patient calibration splits supported intercept-and-slope adjustment. The repeated splits reused the same external patients, and no second cohort tested the adjusted models. The review exercise had no prespecified absolute-risk action threshold and did not assess treatment effects or clinical benefit.

### 4.4 Implications for further evaluation

Larger cohorts with phase-matched CT and consistent lesion definitions could test whether the accuracy pattern persists with better-aligned imaging inputs. Updated models need evaluation in a separate cohort, with probability accuracy and calibration assessed alongside discrimination. Any proposed risk-based review strategy also needs evaluation for its intended clinical use.

## 5 Conclusions

In the cohorts and reference-phase CT representations tested here, adding summaries from one, two, or all retained annotated lesions did not establish an internal first-year survival-risk accuracy advantage over the clinical baseline after TACE. Externally, A-ref had the highest discrimination point estimates, but its discrimination advantage over the clinical baseline remained uncertain and its probability error was greater. Local intercept adjustment reduced A-ref’s median held-out error relative to its original predictions without establishing superiority over adjusted C, and simpler burden features did not improve external accuracy over C. These findings support assessing external probability accuracy and calibration alongside discrimination when comparing lesion representations.

## Data Availability

WAW-TACE is publicly available from Zenodo at https://doi.org/10.5281/zenodo.12741586. HCC-TACE-Seg is publicly available from The Cancer Imaging Archive (TCIA) at https://doi.org/10.7937/TCIA.5FNA-0924. Both datasets are released under the Creative Commons Attribution 4.0 International licence; the WAW-TACE repository additionally specifies use for scientific research. The supplementary material documents cohort assembly, feature definitions, and evaluation methods. The analysis code will be made publicly available upon acceptance. During peer review, the code can be provided to editors and reviewers through the corresponding author.

https://doi.org/10.7937/TCIA.5FNA-0924

## Abbreviations

AFP: alpha-fetoprotein
ALBI: albumin–bilirubin
AP: average precision
AUROC: area under the receiver operating characteristic curve
BCLC: Barcelona Clinic Liver Cancer
CI: confidence interval
CT: computed tomography
HAP: hepatoma arterial-embolization prognostic score
HCC: hepatocellular carcinoma
HU: Hounsfield units
IBS: integrated Brier score
IPCW: inverse probability of censoring weighting
IQR: interquartile range
mHAP-II: modified HAP-II
mRECIST: modified Response Evaluation Criteria in Solid Tumors
NC: noncontrast
PV: portal venous
QC: quality control
TACE: transarterial chemoembolization

## Supplementary information

**Additional file 1 (.pdf): Supplementary methods, tables, and figures**. Tables S1–S10 provide paired comparisons, hyperparameter selections, clinical-score and preprocessing audits, recalibration and prioritization results, cohort differences, complete performance measures, and clinical-variable mapping. Sections S2–S3 document data provenance and detailed methods. Figures S1–S2 show CT–mask quality checks and paired event-capture differences.

## Declarations

### Ethics approval and consent to participate

This study used only publicly available, de-identified data from the WAW-TACE and HCC-TACE-Seg collections. No participants were recruited or contacted, and the authors had no access to identifying information. The original WAW-TACE study was approved by the Bioethics Committee at the Medical University of Warsaw (decision no. AKBE/41/2024) [34]. The original HCC-TACE-Seg study was approved by the Institutional Review Board of The University of Texas MD Anderson Cancer Center, which waived written informed consent because of the retrospective study design [35].

### Consent for publication

Not applicable. The manuscript and supplementary material report aggregate results and non-identifiable CT images. The example images and segmentation overlays were prepared from the publicly released HCC-TACE-Seg collection, distributed under the Creative Commons Attribution 4.0 International licence [40]. The source and licence are identified in the figure caption.

### Availability of data and materials

WAW-TACE is publicly available from Zenodo at https://doi.org/10.5281/zenodo.12741586 [41]. HCC-TACE-Seg is publicly available from The Cancer Imaging Archive (TCIA) [42] at https://doi.org/10.7937/TCIA.5FNA-0924 [40]. Both datasets are released under the Creative Commons Attribution 4.0 International licence; the WAW-TACE repository additionally specifies use for scientific research. Additional file 1 documents cohort assembly, feature definitions, and evaluation methods. The analysis code will be made publicly available upon acceptance. During peer review, the code can be provided to editors and reviewers through the corresponding author.

### Authors’ contributions

ZX and HW designed the study. ZX curated the data, developed the analysis methods and software, conducted the experiments and statistical analyses, validated the results, prepared the visualizations, and wrote the initial draft. AM and EC proofread and revised the manuscript. HW contributed to manuscript writing and revision and supervised the research. All authors reviewed and approved the final manuscript.

## Acknowledgements

We thank the Texas Advanced Computing Center (TACC) at The University of Texas at Austin for providing the computational resources used in this study.

## AI-assisted manuscript preparation

AI tools were used only for language editing and polishing. All authors reviewed and verified the complete manuscript and take responsibility for its accuracy and integrity.

