## Additional file 1: Supplementary methods, tables, and figures for "Lesion selection and external evaluation of clinical–imaging survival models after transarterial chemoembolization for hepatocellular carcinoma"

This supplement accompanies the main article. C denotes the clinical-only model; L1, L2, and A-ref add imaging features from the largest, two largest, or all retained annotated lesions. C+burden adds three tumor-count and volume predictors to C.

### S1 Supplementary Tables

**Table S1.** All paired patient-bootstrap comparisons of the four risk representations.

*Panel A: WAW-TACE*

| Comparison | ΔIBS | $\Delta$AUROC$_{365}$ | ΔC-index |
|---|---|---|---|
| C − L1 | −0.0013 [−0.0072, 0.0046] | −0.0217 [−0.0943, 0.0566] | −0.0148 [−0.0797, 0.0467] |
| C − L2 | 0.0008 [−0.0043, 0.0063] | −0.0176 [−0.0952, 0.0552] | −0.0112 [−0.0780, 0.0508] |
| C − A-ref | −0.0009 [−0.0064, 0.0047] | −0.0192 [−0.0872, 0.0526] | −0.0103 [−0.0718, 0.0485] |
| L1 − L2 | 0.0021 [−0.0009, 0.0056] | 0.0042 [−0.0281, 0.0377] | 0.0035 [−0.0243, 0.0322] |
| L1 − A-ref | 0.0004 [−0.0016, 0.0025] | 0.0025 [−0.0252, 0.0303] | 0.0045 [−0.0202, 0.0289] |
| L2 − A-ref | −0.0017 [−0.0038, 0.0004] | −0.0017 [−0.0233, 0.0195] | 0.0009 [−0.0185, 0.0185] |

*Panel B: HCC-TACE-Seg*

| Comparison | ΔIBS | $\Delta$AUROC$_{365}$ | ΔC-index |
|---|---|---|---|
| C − L1 | −0.0565 [−0.0934, −0.0260] | −0.0121 [−0.1780, 0.1358] | −0.0055 [−0.1640, 0.1295] |
| C − L2 | −0.1020 [−0.1465, −0.0611] | −0.0537 [−0.1974, 0.1007] | −0.0398 [−0.1874, 0.0942] |
| C − A-ref | −0.0645 [−0.0990, −0.0326] | −0.0914 [−0.2448, 0.0429] | −0.0751 [−0.2185, 0.0638] |
| L1 − L2 | −0.0456 [−0.0690, −0.0231] | −0.0416 [−0.1653, 0.0898] | −0.0343 [−0.1546, 0.0810] |
| L1 − A-ref | −0.0080 [−0.0229, 0.0064] | −0.0793 [−0.1542, −0.0095] | −0.0696 [−0.1429, −0.0059] |
| L2 − A-ref | 0.0375 [0.0237, 0.0536] | −0.0377 [−0.1201, 0.0266] | −0.0354 [−0.1089, 0.0308] |

Values are differences [95% CIs], calculated as left representation minus right representation using 2,000 paired patient-level bootstrap replicates. A negative IBS difference favors the left representation because lower IBS is better. A positive AUROC or C-index difference favors the left representation because higher discrimination is better. Internal values use repeated out-of-fold WAW-TACE predictions; external values use coefficient-frozen HCC-TACE-Seg predictions without recalibration.

**Table S2.** Hyperparameter-selection stability in WAW-TACE and missingness of the shared clinical predictors.

*Panel A: Elastic-net selections in repeated nested validation*

| Model | Selected $(C_{\text{reg}}, \rho_{\text{L1}})$: outer fits (out of 15) | Final WAW fit |
|---|---|---|
| C | $(0.01, 0.00)$: 1; $(0.10, 0.00)$: 3; $(0.10, 0.25)$: 6; $(1.00, 0.00)$: 1; $(1.00, 0.25)$: 1; $(1.00, 1.00)$: 3 | $(0.10, 0.00)$ |
| L1 | $(0.01, 0.00)$: 7; $(0.10, 0.00)$: 2; $(0.10, 0.25)$: 1; $(0.10, 0.50)$: 4; $(0.10, 0.75)$: 1 | $(0.01, 0.00)$ |
| L2 | $(0.01, 0.00)$: 1; $(0.10, 0.25)$: 1; $(0.10, 0.50)$: 5; $(0.10, 0.75)$: 3; $(0.10, 1.00)$: 4; $(1.00, 1.00)$: 1 | $(0.10, 0.75)$ |
| A-ref | $(0.01, 0.00)$: 2; $(0.10, 0.25)$: 2; $(0.10, 0.50)$: 5; $(0.10, 0.75)$: 3; $(0.10, 1.00)$: 3 | $(0.10, 0.75)$ |

*Panel B: Missingness of shared clinical predictors*

| Predictor | WAW-TACE ($n = 228$) | HCC-TACE-Seg ($n = 69$) |
|---|---|---|
| Age | 0 | 0 |
| Female sex indicator | 0 | 0 |
| $\log(1 + \text{AFP})$ | 0 | 0 |
| Child–Pugh class | 0 | 0 |
| Ordinal BCLC stage | 0 | 0 |

Panel A counts the 15 outer fits per representation (three repetitions of five-fold cross-validation); each fit contributes one selected hyperparameter pair. Final-fit hyperparameters were selected on all 228 WAW-TACE patients before frozen transport to HCC-TACE-Seg. Panel B reports counts missing before imputation; cohort denominators are shown in the headings. BCLC, Barcelona Clinic Liver Cancer.

**Table S3.** Audit of published clinical-score reconstructability.

| Score | Proxy fields checked | WAW with all proxies | HCC with all proxies | Exact reconstruction |
|---|---|---|---|---|
| ALBI | Albumin; bilirubin | 228/228 | 0/69 | No |
| HAP | Albumin; bilirubin; $\log(1 + \mathrm{AFP})$; largest-lesion volume | 228/228 | 0/69 | No |
| mHAP-II | Albumin; bilirubin; $\log(1 + \mathrm{AFP})$; largest-lesion volume; total annotated lesion count | 228/228 | 0/69 | No |
| Six-and-Twelve | Total annotated lesion count; largest-lesion three-dimensional bounding-box diagonal | 228/228 | 69/69 | No |

Counts indicate patients with all listed proxy fields present, not patients with verified inputs for the published score. Albumin and bilirubin were unavailable in the HCC analytic inputs. WAW proxy completeness did not verify source-compatible score definitions and units. For HAP, mHAP-II, and Six-and-Twelve, lesion volume or a three-dimensional bounding-box diagonal did not provide the required diameter measurement. None of the four scores was calculated or approximated.

**Table S4.** Outcome-blind lesion, geometry, and reference-frame quality-control audit.

| Audit item | WAW-TACE | HCC-TACE-Seg |
|---|---|---|
| Patients in the source cohort | 233 | 105 |
| Patients assessed for this analysis | 233 | 69 |
| Model-eligible patients | 228 | 69 |
| Indexed CT scans | 854 | – |
| Available phases (NC/A/PV/D) | 200/230/231/193 | 69/69/69/– |
| Reference image | Mask-associated CT phase | Pretreatment PV |
| Mask-associated reference phases before lesion QC (NC/A/PV/D) | 3/126/74/30 | –/–/69/– |
| Lesion instances/components before image QC | 377 | 122 |
| Retained lesion instances/components | 372 | 122 |
| Non-retained lesion instances/components | 5 | 0 |
| Outcome-blind geometry exclusions | 1 | 0 |
| Reference-grid QC failures | 0 | 0 |
| Patients missing any L1/L2/A-ref representation | 0 | 0 |
| Model-eligible patients with multiple retained annotated lesions | 85 | 14 |
| Sampled three-slice CT–mask review | – | 11 reviewed |
| Missing CT phases imputed | No | No |
| Survival outcome used for component or reference QC | No | No |

NC, noncontrast/native; A, late arterial; PV, portal venous; D, delayed. WAW scan and phase counts refer to the 233 assessed patients before lesion QC; HCC phase counts refer to the 69-patient triphasic subset. Representation availability and multiple-lesion counts refer to the model-eligible cohorts (228 WAW and 69 HCC patients). Dashes denote quantities not tabulated in this audit. No gross CT–mask misalignment was identified in three-slice views of the 11 sampled HCC patients (Figure S1). The frozen audit found no duplicate instance keys or retained lesion identifiers, nonpositive retained volumes, nonfinite reference-lesion features, or invalid overall-survival records after the endpoint join. The five WAW exclusions involved one annotated lesion per patient: one had unresolved CT–mask geometry and four did not meet the 80% liver-support threshold.

During assembly of the HCC subset, 34 of 105 patients did not meet the image-matching criteria. In 27 patients, the CT series referenced by the tumor segmentation could not be directly matched to an author-curated CT series. No qualifying NC series was identified in five patients and no qualifying A series in one. In one further patient, the segmentation referenced an A rather than a PV series. Of the remaining 71 patients, one was excluded for irregular PV slice spacing and one because tumor-mask conversion failed, leaving 69 patients.

**Table S5.** Local recalibration feasibility and held-out performance in HCC-TACE-Seg.

*Panel A: Within-model changes relative to original predictions*

| Model | Adjustment | Calibration patients | Supported splits out of 200 | $\Delta$Brier median [Q1, Q3] |
|---|---|---|---|---|
| C | Intercept | 10 | 6 | $0.0542$ [$0.0416$, $0.0796$] |
| C | Intercept | 20 | 84 | $-0.0052$ [$-0.0071$, $0.0026$] |
| C | Intercept | 40 | 200 | $-0.0061$ [$-0.0088$, $0.0004$] |
| C | Intercept and slope | 10 | 0 | – |
| C | Intercept and slope | 20 | 0 | – |
| C | Intercept and slope | 40 | 67 | $-0.0044$ [$-0.0077$, $0.0027$] |
| A-ref | Intercept | 10 | 6 | $-0.0199$ [$-0.0258$, $-0.0161$] |
| A-ref | Intercept | 20 | 84 | $-0.0598$ [$-0.0643$, $-0.0541$] |
| A-ref | Intercept | 40 | 200 | $-0.0616$ [$-0.0794$, $-0.0361$] |
| A-ref | Intercept and slope | 10 | 0 | – |
| A-ref | Intercept and slope | 20 | 0 | – |
| A-ref | Intercept and slope | 40 | 67 | $-0.0880$ [$-0.1017$, $-0.0759$] |

*Panel B: Adjusted A-ref versus adjusted C on identical held-out patients*

| Adjustment | Calibration patients | Matched splits | Adjusted A-ref $-$ C median [Q1, Q3] | Splits favoring A-ref ($n/N$) |
|---|---|---|---|---|
| Intercept | 10 | 6 | 0.0129 [0.0006, 0.0198] | 2/6 |
| Intercept | 20 | 84 | 0.0225 [0.0131, 0.0324] | 8/84 |
| Intercept | 40 | 200 | 0.0204 [0.0031, 0.0366] | 41/200 |
| Intercept and slope | 40 | 67 | 0.0071 [$-0.0066$, 0.0155] | 24/67 |

Panel A reports adjusted minus original Brier score within each split; panel B subtracts adjusted C from adjusted A-ref on the same held-out patients. Negative values indicate lower error for adjustment in A and for A-ref in B. Both models had identical supported splits. Only these splits contribute performance summaries, so adjustment methods with different support counts are not compared on the full set of 200 splits. Intercept adjustment required at least five deaths and five known survivors in the calibration subset; intercept-and-slope adjustment required at least 10 of each. Brackets show Q1 and Q3 across overlapping splits, not confidence limits; dashes indicate that no supported splits were available. Only day-365 risk was recalibrated; adjusted 0–365-day IBS was not estimated.

**Table S6.** Day-365 death capture at fixed review capacities in HCC-TACE-Seg.

| Model | Review capacity | Selected $n$ | Observed deaths | IPCW capture % [95% CI] | IPCW event proportion (%) |
|---|---|---|---|---|---|
| C | 10% | 7 | 3 | 20.0 [5.6, 43.5] | 42.7 |
| A-ref | 10% | 7 | 3 | 20.0 [5.8, 41.7] | 42.7 |
| C | 20% | 14 | 7 | 46.5 [21.3, 66.5] | 49.8 |
| A-ref | 20% | 14 | 5 | 33.3 [16.5, 64.0] | 35.6 |
| C | 30% | 21 | 8 | 53.1 [30.6, 78.6] | 37.9 |
| A-ref | 30% | 21 | 8 | 53.2 [30.0, 78.1] | 37.9 |

Models selected the same number of patients using original, uncalibrated HCC predictions. Observed deaths are raw counts among selected patients. IPCW capture is the weighted fraction of all cohort deaths captured; IPCW event proportion is the weighted death proportion within the selected patients. Both IPCW measures are percentages. There were 15 observed deaths by day 365 and one patient censored before that day. Brackets show 95% CIs from 2,000 patient-level bootstrap replicates; paired model differences are shown in Figure S2. This retrospective ranking exercise does not estimate the effect of further clinical assessment.

**Table S7.** Paired performance comparisons for the additional C+burden model.

*Panel A: Probability accuracy*

| Cohort | Comparison | ΔIBS | $\Delta\text{Brier}_{365}$ |
|---|---|---|---|
| WAW-TACE | C+burden − C | 0.0002 [−0.0053, 0.0062] | −0.0104 [−0.0201, −0.0011] |
| | C+burden − A-ref | −0.0007 [−0.0038, 0.0026] | −0.0039 [−0.0106, 0.0023] |
| HCC-TACE-Seg | C+burden − C | 0.0283 [0.0012, 0.0606] | 0.0477 [−0.0016, 0.1019] |
| | C+burden − A-ref | −0.0361 [−0.0588, −0.0152] | −0.0256 [−0.0647, 0.0181] |

*Panel B: Discrimination*

| Cohort | Comparison | $\Delta\text{AUROC}_{365}$ | ΔC-index |
|---|---|---|---|
| WAW-TACE | C+burden − C | 0.0408 [−0.0175, 0.0952] | 0.0296 [−0.0162, 0.0821] |
| | C+burden − A-ref | 0.0215 [−0.0211, 0.0606] | 0.0193 [−0.0129, 0.0504] |
| HCC-TACE-Seg | C+burden − C | −0.0322 [−0.1722, 0.1449] | −0.0331 [−0.1679, 0.1456] |
| | C+burden − A-ref | −0.1236 [−0.2300, −0.0246] | −0.1083 [−0.2150, −0.0210] |

Values are differences [95% CIs], calculated as left model minus right model using 1,000 paired patient-level bootstrap replicates. Negative IBS and Brier differences favor the left model; positive AUROC and C-index differences favor the left model. C+burden was trained by the original WAW nested procedure and transported to HCC without refitting or recalibration.

**Table S8.** Analytic-cohort characteristics and selected outcome-blind predictor differences.

**Panel A: Shared clinical predictors**

| Characteristic | WAW-TACE ($n = 228$) | HCC-TACE-Seg ($n = 69$) |
|---|---|---|
| Age, years | 66 [60, 73] | 71 [60, 76] |
| Female sex, $n$ (%) | 48 (21.1) | 30 (43.5) |
| $\log(1 + \text{AFP})$, AFP measured in ng/mL | 2.76 [1.81, 5.71] | 3.99 [1.97, 7.31] |
| Child–Pugh class A, $n$ (%) | 202 (88.6) | 58 (84.1) |
| Child–Pugh class B, $n$ (%) | 26 (11.4) | 11 (15.9) |
| Child–Pugh class C, $n$ (%) | 0 (0.0) | 0 (0.0) |
| BCLC stage 0/A, $n$ (%) | 165 (72.4) | 6 (8.7) |
| BCLC stage B, $n$ (%) | 63 (27.6) | 17 (24.6) |
| BCLC stage C, $n$ (%) | 0 (0.0) | 44 (63.8) |
| BCLC stage D, $n$ (%) | 0 (0.0) | 2 (2.9) |

**Panel B: Selected predictor shifts**

| Feature | WAW median | HCC median | Standardized mean difference |
|---|---|---|---|
| Ordinal BCLC stage | 0 | 2 | 2.289 |
| Annotated lesion count | 1 | 1 | 0.082 |
| Total tumor volume, mL | 50.1 | 141.2 | 0.577 |
| Largest-lesion volume, mL | 45.1 | 141.2 | 0.577 |
| Largest-lesion bounding-box diagonal, mm | 81.9 | 123.3 | 0.807 |
| Largest-lesion sphericity | 0.642 | 0.522 | −1.990 |
| Largest-lesion relative attenuation, HU | 7 | −10 | −0.628 |

Panel A reports median [Q1, Q3] for continuous predictors and $n$ (%) for categorical predictors. All five shared clinical predictors were available for every patient in both analytic cohorts. Panel B reports the standardized mean difference, $(\bar{x}_{\text{HCC}} - \bar{x}_{\text{WAW}})/\sqrt{(s^2_{\text{HCC}} + s^2_{\text{WAW}})/2}$, where $\bar{x}$ is the cohort mean and $s^2$ the sample variance. Positive values indicate a higher HCC mean; displayed medians are not used in the calculation. These outcome-blind summaries are descriptive. The model-eligible WAW reference phases were arterial (123), portal venous (72), delayed (30), and native (3); all 69 HCC reference images were portal venous.

**Table S9.** Complete internal and frozen external performance measures.

*Panel A: WAW-TACE repeated nested out-of-fold predictions*

| Measure | C | L1 | L2 | A-ref | C+burden |
|---|---|---|---|---|---|
| $\text{Brier}_{365}$ | 0.1842 | 0.1797 | 0.1751 | 0.1778 | 0.1738 |
| $\text{AUROC}_{365}$ | 0.6529 | 0.6746 | 0.6704 | 0.6721 | 0.6937 |
| $\text{AP}_{365}$ | 0.4200 | 0.4452 | 0.4934 | 0.4608 | – |
| IBS | 0.0942 | 0.0955 | 0.0934 | 0.0951 | 0.0944 |
| C-index | 0.6337 | 0.6484 | 0.6449 | 0.6440 | 0.6632 |
| Calibration intercept | −0.257 | −0.053 | 0.094 | 0.047 | 0.161 |
| Calibration slope | 0.744 | 0.929 | 1.073 | 1.041 | 1.148 |

*Panel B: HCC-TACE-Seg coefficient-frozen external predictions*

| Measure | C | L1 | L2 | A-ref | C+burden |
|---|---|---|---|---|---|
| $\text{Brier}_{365}$ | 0.1583 | 0.2342 | 0.2749 | 0.2317 | 0.2060 |
| $\text{AUROC}_{365}$ | 0.6351 | 0.6472 | 0.6889 | 0.7265 | 0.6029 |
| $\text{AP}_{365}$ | 0.4780 | 0.3287 | 0.3747 | 0.4312 | – |
| IBS | 0.0631 | 0.1195 | 0.1651 | 0.1276 | 0.0914 |
| C-index | 0.6320 | 0.6376 | 0.6718 | 0.7072 | 0.5989 |
| Calibration intercept | −0.530 | −1.254 | −1.474 | −1.371 | −1.190 |
| Calibration slope | 0.886 | 0.245 | 0.315 | 0.467 | 0.138 |

AP, average precision; IBS, integrated Brier score over 0–365 days. The calibration intercept and slope were estimated jointly. Brier score, AUROC, AP, and calibration coefficients are reported at day 365; C-index uses day-365 risk and comparable pairs with an earlier death by day 365. These calibration coefficients did not modify the primary predictions. IBS confidence intervals are reported in Table 3 of the main article. C+burden was evaluated separately after the primary four-model analysis; its AP was not reported (–).

### S2 Data Provenance and Analysis Records

The source cohorts and annotations are described in the WAW-TACE and HCC-TACE-Seg data publications [1, 2]. Preprocessing and modeling outputs were stored in versioned directories without overwriting source images or masks. The study record includes endpoint mappings, SHA-256 file checksums (content fingerprints used to identify input-file versions), WAW mask-associated CT phase assignments, lesion-instance and reference-grid checks, feature definitions, patient-level splits, model settings, out-of-fold and external predictions, hyperparameter selections, and fitted coefficients.

### S3 Supplementary Methods

#### S3.1 Clinical-variable mapping

Table S10 maps source fields to the five shared clinical predictors. Numeric fields were parsed before modeling. Missing or unparseable values remained missing for training-set median imputation. AFP values below zero were treated as missing rather than log-transformed. Child–Pugh and BCLC were ordered numeric inputs, not one-hot indicators. All five predictors, including binary sex, underwent the training-set standardization described in Section 2.3 of the main article.

#### S3.2 Lesion geometry and attenuation

Masks were reconciled to the reference CT grid only when spatial metadata described an equivalent physical coordinate frame. Identity resampling was recorded as grid reconciliation rather than image registration. Physical lesion volume was the mask voxel count multiplied by the voxel volume. Bounding-box extent on each axis included the full span of occupied voxels and was converted to millimeters using image spacing; the diagonal was the Euclidean norm of these three extents. Surface area $A$ was estimated by summing exposed voxel-face areas. With volume $V$ in $\text{mm}^3$, surface-to-volume ratio was $A/V$ and sphericity was $\pi^{1/3}(6V)^{2/3}/A$. These are voxel-based geometric descriptors, not manually measured diameters.

**Table S10.** Source-field mapping for the five shared clinical predictors.

| Predictor | WAW-TACE field | HCC-TACE-Seg field | Shared model encoding |
|---|---|---|---|
| Age | age | age | Numeric age in years. |
| Sex | gender_woman | Sex | Male = 0, female = 1. WAW codes retained; HCC codes 1 and 2 mapped to 0 and 1. |
| AFP | lab_afp | AFP | $\log(1 + \mathrm{AFP})$, with AFP in ng/mL. |
| Child–Pugh class | cps | CPS | Classes A/B/C = 1/2/3. WAW numeric class codes retained; HCC class labels mapped. |
| BCLC stage | bclc | BCLC | Stages 0/A, B, C, D = 0, 1, 2, 3. WAW codes 0/1 retained; HCC labels mapped, grouping stage 0 with A. |

*Note.* Field names reproduce the source workbooks. The Child–Pugh predictor represents class, not the 5–15-point score. Unrecognized HCC sex or class/stage labels and WAW BCLC codes outside 0/1 were treated as missing. Source endpoint fields were kept separate from predictors; HCC survival time in weeks was converted to days by multiplying by seven.

CT values were clipped before computing attenuation features. For each lesion, the reference liver region comprised voxels in the provided liver mask after excluding that lesion. Median lesion HU and median HU in this reference region were used to calculate relative attenuation and the high-attenuation fraction in Table 2 of the main article. Surface area and these two medians were retained as intermediate audit values, but were not separate model inputs. For spatial-support QC, WAW used the provided liver segment. HCC used the union of liver and lesion masks because its source segments were mass-exclusive; the support rule and raw liver overlap were recorded separately. This HCC support rule should not be interpreted as an independent test that a lesion lies inside the raw liver segment.

Lesions were ordered by decreasing physical volume. Equal-volume lesions were ordered by their physical centroid coordinates $(x, y, z)$, followed by source mask path in WAW or component label in HCC. For a selected set of $m$ lesions with feature values $f_j$ and volumes $v_j$, the volume-weighted mean was $\sum_j v_j f_j / \sum_j v_j$. The standard deviation was unweighted, with divisor $m$, and the range was $\max_j f_j - \min_j f_j$. No additional lesion was supplied when a patient had fewer than two lesions. With one selected lesion, the mean and maximum equal its value, while the standard deviation and range are zero. The selected tumor volume and small-lesion volume fraction were computed only from selected lesions; total annotated lesion count used all retained lesions. This count was not a clinical workbook's reported tumor count.

#### S3.3 Pooled-logistic design and penalized likelihood

Let $\mathcal{O}$ denote the set of included patient–interval records and $y_{ik}$ their binary death labels. The four intervals were $(0, 91]$, $(91, 182]$, $(182, 273]$, and $(273, 365]$ days. A death contributed its containing interval and any completed earlier intervals. A censored patient contributed only intervals whose end was no later than the censoring time. An incomplete censored interval did not contribute a negative label or a fractional record.

After patient-level preprocessing, the design row was

$$\mathbf{z}_{ik} = \left(\mathbf{x}_i^\top, \mathbf{1}\{k = 2\}, \mathbf{1}\{k = 3\}, \mathbf{1}\{k = 4\}\right)^\top .$$

Here, $\mathbf{1}\{\cdot\}$ is 1 when its condition is true and 0 otherwise. Writing $\alpha_k = a + d_k$, with $d_1 = 0$, gives $\boldsymbol{\theta} = (\boldsymbol{\beta}^\top, d_2, d_3, d_4)^\top$ and linear predictor $a + \boldsymbol{\theta}^\top \mathbf{z}_{ik}$. The likelihood term in Section 2.3 of the main article was

$$\mathcal{L}(a, \boldsymbol{\theta}) = - \sum_{(i,k)\in\mathcal{O}} \big[ y_{ik} \log h_i(k) + (1 - y_{ik}) \log\{1 - h_i(k)\} \big] .$$

The L1 norm is the sum of absolute coefficients and the squared L2 norm is the sum of squared coefficients. The elastic-net penalty applied to all entries of $\boldsymbol{\theta}$, but not to $a$. The three interval indicators were appended after standardizing patient predictors and were not themselves standardized. Thus, the four interval intercepts were learned rather than supplied as patient inputs.

Elastic-net models used the scikit-learn SAGA solver with a tolerance of $10^{-4}$ and a maximum of 10,000 iterations. Cross-validation folds were stratified by the observed death indicator. The outer-partition seeds were 20260908, 20260909, and 20260910; the final hyperparameter selection used seed 20260912. Ties in mean inner-validation IBS were resolved by smaller $C_{\mathrm{reg}}$, then smaller $\rho_{\mathrm{L1}}$.

#### S3.4 Numerical integration, calibration, and uncertainty

IBS summarizes IPCW Brier scores over follow-up [3, 4] and was approximated by the trapezoidal rule on $t = (0, 91, 182, 273, 365)$ days, with Brier score at day 0 set to zero:

$$\widehat{\mathrm{IBS}} = \frac{1}{365} \sum_{j=1}^{4} \frac{t_j - t_{j-1}}{2} \left\{ \widehat{\mathrm{BS}}(t_{j-1}) + \widehat{\mathrm{BS}}(t_j) \right\}.$$

Let $T_i$ denote the observed death or censoring time, $\delta_i$ the death indicator, and $\widehat{G}$ the Kaplan–Meier estimate of censoring survival in the evaluated sample [5]. At horizon $t$, patients who died by $t$ received weight $1/\widehat{G}(T_i^-)$, and patients observed beyond $t$ received weight $1/\widehat{G}(t)$; other patients received zero weight. Here, $T_i^-$ denotes the instant immediately before the observed death time. The denominator in each IPCW Brier score was the evaluated cohort size $n$, not the sum of weights. When calculating weights, $\widehat{G}$ was bounded below by $10^{-6}$. Censoring probabilities were estimated separately from predictor preprocessing and model fitting. These marginal Kaplan–Meier weights assume that censoring is independent of event time and the predictors within the evaluated sample. Covariate-dependent censoring was not modeled.

Time-dependent AUROC and average precision used the observable binary outcomes and the same IPCW weights defined above.

Harrell's C-index [6] used the original predicted day-365 risk. A pair was comparable when one patient died by day 365 and the other had a strictly later observed death or censoring time. A higher predicted risk for the earlier death scored 1, a tied risk scored 0.5, and a lower risk scored 0. Pairs with equal observed times were excluded. The index was the mean score over comparable pairs and did not use IPCW.

Calibration intercept $a_{\mathrm{cal}}$ and slope $b_{\mathrm{cal}}$ were estimated jointly by an IPCW-weighted logistic regression of observable 365-day death status on logit predicted risk [7]:

$$\mathrm{logit}\{p_{i,\mathrm{cal}}\} = a_{\mathrm{cal}} + b_{\mathrm{cal}}\, \mathrm{logit}\{\widehat{R}_i(365)\}.$$

The jointly estimated intercept is the intercept of this calibration model, rather than a calibration-in-the-large estimate obtained with the slope fixed at 1. Predictions were clipped to $[10^{-5}, 1 - 10^{-5}]$ for this calculation. Ideal calibration corresponds to $(a_{\mathrm{cal}}, b_{\mathrm{cal}}) = (0, 1)$. These diagnostic coefficients did not change the primary predictions.

Bootstrap replicates resampled whole patients and their prediction trajectories, preserving pairing across models. Confidence limits were the 2.5th and 97.5th percentiles of replicate estimates. Models were not retrained within these evaluation bootstraps; the intervals summarize resampling uncertainty for the reported prediction sets, not variability from repeating the complete development procedure.

#### S4 Supplementary Figures

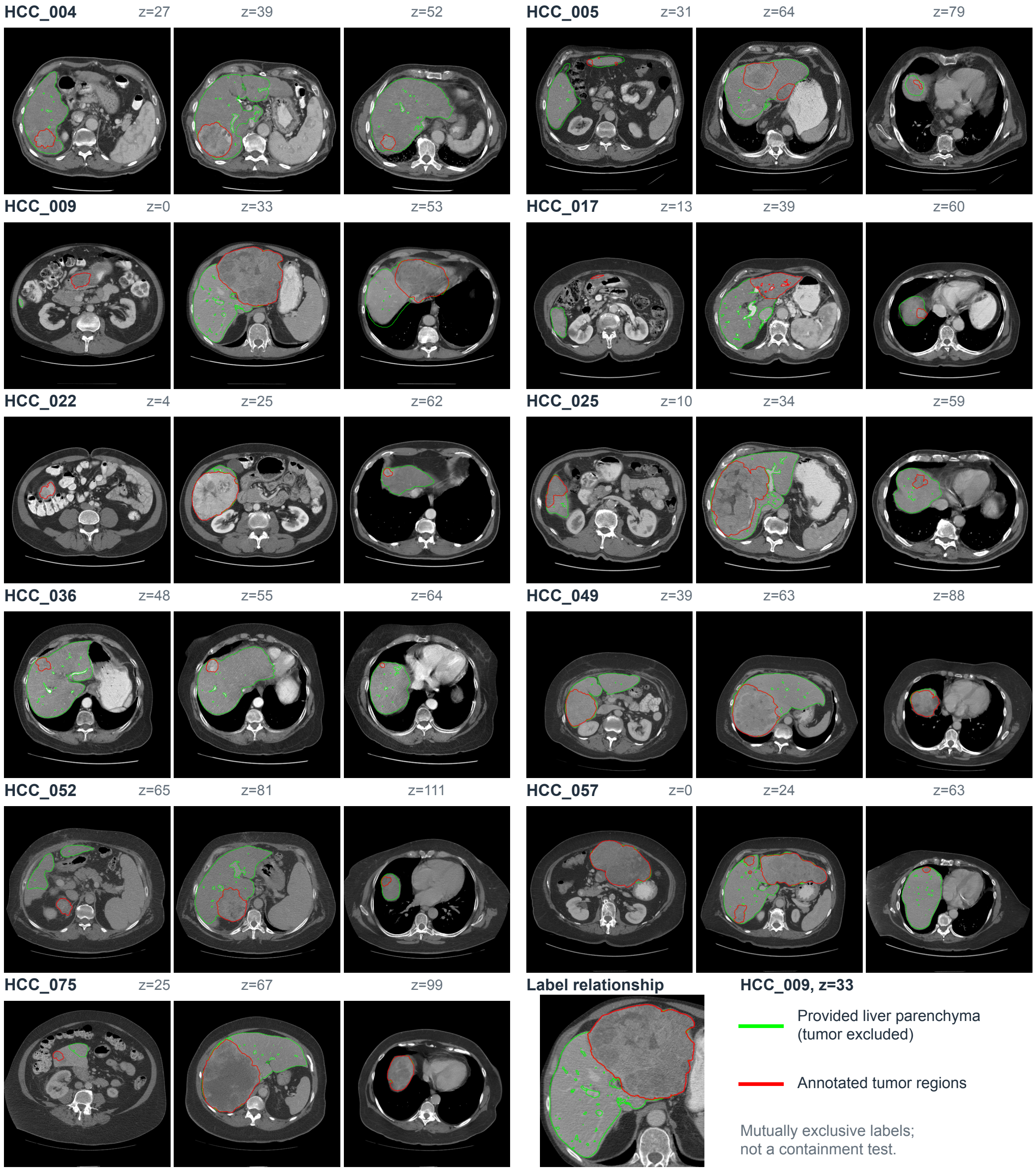


**Figure S1.** Visual checks of CT–mask alignment in an outcome-blind sample of 11 HCC-TACE-Seg patients. Three axial slices per patient span the tumor annotations. Green outlines show the provided liver-parenchyma segment (tumor excluded); red outlines show tumor annotations. These segments are mutually exclusive, not nested. The inset enlarges HCC_009 at axial slice index 33. The sample included multiple-component, small-component, and boundary cases. No gross CT–mask misalignment was identified in the sampled views. CT images and segmentations are from HCC-TACE-Seg [8], available under CC BY 4.0; slice selection, coloured overlays, and the enlarged inset were added for this figure.

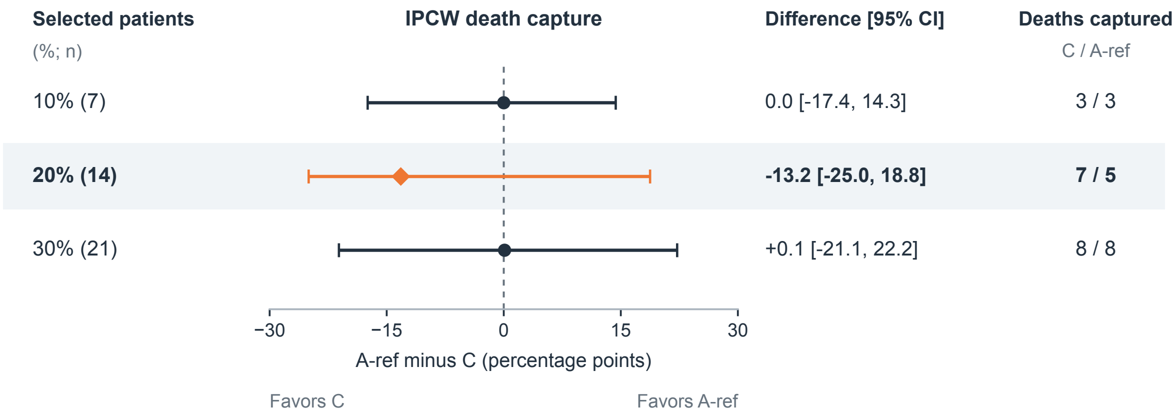


**Figure S2.** Paired A-ref-minus-C differences in day-365 death capture in HCC-TACE-Seg. IPCW estimates use the original, uncalibrated frozen predictions. Intervals are 95% confidence intervals from 2,000 paired patient-bootstrap resamples. Shading marks the prespecified 20% review capacity; 10% and 30% are sensitivity analyses. Death counts are unweighted. Differences are in percentage points; negative values favor C. This retrospective ranking analysis does not estimate clinical benefit.
